# Distinct Global Patterns and Trends in Lifetime Risk of Rectal Cancer Within Colorectal Cancer: A Population-Based Analysis from GLOBOCAN 2022

**DOI:** 10.64898/2026.03.30.26349699

**Authors:** Ke Pang, Xiyi An, Kai Song, Feng Xie, Hanyue Ding, Haitao Zhou, Zhonghu He, Hongda Chen, Dong Wu

## Abstract

**Background:** Rectal cancer (RC) is traditionally grouped within colorectal cancer (CRC), despite growing evidence of distinct epidemiologic features. However, global comparative assessments of lifetime risks of RC relative to CRC remain limited. We aimed to estimate lifetime risks of developing and dying from RC and CRC worldwide and to examine geographic, socioeconomic, and temporal variations in the proportional contribution of RC within CRC.

**Methods:** Age-specific incidence and mortality estimates for RC and CRC across 185 countries were obtained from GLOBOCAN 2022, together with population and all-cause mortality data from the United Nations. Lifetime risks of incidence (LRI) and mortality (LRM) were calculated using the adjusted-for-multiple-primaries (AMP) method by sex, country, region, and Human Development Index (HDI). The RC-to-CRC lifetime risk ratio quantified the proportional contribution of RC. Temporal trends were assessed in 42 countries using Cancer Incidence in Five Continents Plus (CI5plus) data and average annual percent change (AAPC).

**Results:** In 2022, the global lifetime risk of developing RC was 1.61% and dying from RC was 0.95%, accounting for approximately 35% of the corresponding CRC lifetime burden (4.61% and 2.68%). Absolute lifetime risks of both RC and CRC increased with HDI. In contrast, the proportional contribution of RC varied markedly, peaking at 41%–43% in Central and South-Eastern Asia but falling below 20% in the Caribbean and Central America, and showed a negative association with HDI. The LRI/LRM ratio increased with socioeconomic development. Temporal analyses showed increasing LRI trends in 17 of 42 countries for CRC versus 9 for RC, while declines occurred in 14 countries for RC and 11 for CRC.

**Conclusions:** RC constitutes a substantial yet epidemiologically distinct component of the global CRC burden. Its proportional contribution varies across regions and does not parallel absolute risk patterns, supporting the need for subsite-specific surveillance and prevention strategies.

## Introduction

Colorectal cancer (CRC) remains one of the most prevalent malignancies worldwide, ranking as the third most diagnosed cancer and the second leading cause of cancer-related death^[1]^. Although incidence has stabilized or declined in many high-income regions due to improved screening and management, the burden continues to rise rapidly in newly industrialized and medium-human development index (HDI) countries, particularly across Asia, Eastern Europe, and Latin America ^[2]^. These divergent trajectories underscore substantial global heterogeneity in CRC epidemiology.

Within CRC, age- and region-specific shifts have become increasingly apparent. Rectal cancer (RC) incidence has risen among younger adults in Western countries—including the United States, Canada, and Australia—despite decreasing trends in older populations, whereas the overall CRC burden continues to increase in many transitioning regions^[3]^. Such contrasting patterns suggest underlying etiologic differences between subsites of the large bowel. Supporting this, marked geographical variation is observed in subsite distribution: RC accounts for <40% of CRCs in most European and North American countries but exceeds 50% across many Asian populations, including China^[4]^. The reasons remain unclear but imply region-specific exposures and warrant closer investigation.

Environmental and lifestyle factors appear to contribute differentially to colon and rectal carcinogenesis. In lower-HDI settings, particularly rural Asia, environmental pollution, including contaminated surface water and chlorination by-products (e.g., chloroform, carbon tetrachloride), and agricultural exposure to organophosphorus and chlorine-containing pesticides have been implicated in elevated RC risk^[5–7]^. By contrast, in high-income regions, high-fat diets, red meat intake, and physical inactivity are more strongly associated with colon cancer, whereas alcohol consumption and smoking exert greater effects on RC^[3]^. Findings on diet further support this heterogeneity: ultra-processed foods are associated with colon cancer but not RC, whereas dietary patterns rich in red/processed meats and alcohol are linked to RC but not proximal colon tumors^[8,9]^. Females who smoke may have a higher risk of RC due to smoking than their male counterparts^[10]^.

Genetic and molecular evidence reinforces this divergence. Familial aggregation studies show higher heritability for colon than rectal cancer, with site-specific associations with extracolonic malignancies—lung cancer for RC and nervous system cancers for colon cancer^[11,12]^. Mechanistically, colon cancers more commonly harbor mismatch repair deficiency and APC mutations, whereas RCs demonstrate overexpression of COX2 and prostaglandin E□. Large-scale genomic analyses reveal distinct chromosomal alterations, mutational spectra, and transcriptomic programs, including WNT/MYC/mTOR pathway dominance in colon cancer and GPCR/neuronal pathway enrichment in RC^[13]^. RCs additionally exhibit unique transcriptional subtypes (RSS1–RSS3) with differential immune infiltration, EMT activity, and therapeutic responsiveness^[14]^. Metabolomic studies further identify distinct sphingolipid landscapes, underscoring metabolic divergence.

Despite accumulating evidence that RC constitutes a distinct disease entity, global cancer statistics still aggregate colon and RCs into a single category, masking important site-specific disparities in burden, risk, and outcomes. Lifetime risk estimation offers an integrative measure of population susceptibility, combining age-specific incidence with competing mortality risks. It is also more intuitive for clinicians, policymakers, and the public, because it can be interpreted as the probability that an individual will develop or die from a disease over the remaining lifespan. This feature may be particularly valuable for international comparisons across countries with markedly different life expectancy profiles, where crude or age-standardized rates may not fully capture cumulative population risk.

In this study, we systematically estimated and compared the lifetime risk of colorectal cancers across countries and HDI strata using the adjusted-for-multiple-primaries (AMP) method. We further examined the proportion of RC within total CRC, residual lifetime risks at ages 50, 60, and 70, and temporal trends using average annual percent change (AAPC). By elucidating global variations and socioeconomic inequalities in colon and RC risks, our findings aim to support site-specific prevention strategies and strengthen the rationale for a distinct research framework for RC.

## Methods

### Data Sources

The data for new cancer cases, deaths, and population by sex and age group (0-4, 5-9, …, 65-69, 70-84, and 85+) in 185 countries were sourced from GLOBOCAN 2022, which offers the latest global cancer burden estimates developed by the International Agency for Research on Cancer (IARC). Detailed descriptions of the data sources and methods for compiling the GLOBOCAN estimates can be found on the Global Cancer Observatory (GCO) website (https://gco.iarc.who.int). Briefly, national estimates for different cancer types are based on the best available data on cancer incidence and mortality from each country. These estimates primarily utilize short-term projections and modeled mortality-to-incidence ratios, and are presented by country, region, sex, and age group. Cancer types were classified following the tenth edition of the International Classification of Diseases (ICD-10). We extracted data for RC (ICD-10: C19–C20) and CRC (ICD-10: C18–C21).

Along with cancer type data, we also obtained corresponding death rates and population statistics. The population and all-cause mortality data were provided by the United Nations (World Population Prospects 2019, (https://population.un.org/wpp/).

To estimate cancer deaths, we applied the mortality-to-incidence ratios from GLOBOCAN 2022 to the case counts from CI5 Plus, stratified by cancer type, age group, sex, and year. All-cause deaths were calculated by applying United Nations all-cause mortality rates to the population data from CI5 Plus, also stratified by age group, sex, and year. Additionally, we incorporated HDI data from the United Nations Development Programme (https://hdr.undp.org/), which measures the socioeconomic development of a country or region through indicators such as life expectancy, literacy, and gross domestic product (GDP) per capita.

For trend analysis, cancer registry data from the Cancer Incidence in Five Continents (CI5) Plus database (https://ci5.iarc.fr/ci5plus/) were used, covering the years 2003 to 2017. The methodology and quality control standards applied in the CI5 Plus database are consistent with the CI5 series, which underpins GLOBOCAN estimates. All registries included in the database meet IARC’s quality control criteria, ensuring data comparability, completeness, validity, and timeliness. After excluding countries with incomplete data for certain age groups or fewer than 10 reported cases annually for each cancer type, 42 countries with complete data for all three cancer sites were included in the trend analysis.

### Statistical methods

We estimated the lifetime risk of RC and CRC using the adjusted-for-multiple-primaries (AMP) method. The technical details of the AMP algorithm have been described previously and are provided in the **Supplementary Material**. In brief, the AMP method incorporates age-specific all-cause mortality, disease-specific incidence, and disease-specific mortality, categorized into 5-year age groups, to cumulatively compute the lifetime risk of developing a disease (LRI) and the lifetime risk of dying from a disease (LRM). These indicators represent the probability that an individual from a given population will develop or die from colon or RC during their remaining lifetime, conditional on survival to the starting age.

We calculated the LRI and LRM for RC and overall CRC across 185 countries, stratified by sex (both sexes, male, female) and selected age intervals, including birth to death, as well as residual lifetime risks at ages 50, 60, and 70 years.

To further characterize the anatomical distribution of CRC burden, we additionally computed the proportion of rectal cancer within total CRC lifetime risk (RC-to-CRC LRI ratio and RC-to-CRC LRM ratio). This indicator was evaluated in parallel with CC-specific and RC-specific lifetime risks across all analyses, including by country, by HDI category, and by age interval.

Countries were grouped into four Human Development Index (HDI) categories (very high, high, medium, and low). Lifetime risks for each HDI category were obtained by pooling country-level data using population-weighted aggregation. In addition, we classified the world into 20 predefined geographic regions, and estimated regional lifetime risks of CC, RC, and RC-to-CRC proportions by aggregating the constituent countries within each region.

To further examine the relative contributions of incident risk and mortality burden, we calculated the LRI-to-LRM ratio for CC and RC separately. To assess the contribution of older-age onset, we calculated the ratio of residual lifetime risks at age 60 to lifetime risks from birth (60-to-whole risk proportion).

Temporal trends in lifetime risks were evaluated by estimating the average annual percent change (AAPC) using log-linear joinpoint regression for countries with ≥15 years of consecutive cancer registry data. Separate AAPCs were computed for CC, RC, and the RC-to-CRC proportion to identify site-specific and region-specific temporal patterns.

All analyses were performed using R version 4.5.0, following the methodological framework detailed in the Supplementary Appendix.

## Results

### Global lifetime risks of colorectal malignancies

In 2022, the global lifetime risk of developing RC was 1.61% (95% CI: 1.61–1.62), accounting for approximately 35% of all lifetime CRC risk, which reached 4.61% (4.61–4.62). A similar proportion was observed for the lifetime risk of dying, with RC representing 35% of CRC deaths globally (0.95% vs. 2.68%). Marked sex differences were observed. The lifetime risk of developing RC was 1.99% in males and 1.28% in females, corresponding to 38% and 32% of their respective lifetime CRC risks. The lifetime risk of RC mortality showed an even greater male predominance (1.29% in males vs. 0.75% in females), representing 42% and 32% of CRC lifetime mortality, respectively.

### Lifetime risks by geographical region

Regionally, the lifetime risk of developing RC varied substantially, ranging from the highest in Northern Europe (2.76% in both sexes combined) and Australia/New Zealand (2.73%) to the lowest in Western Africa (0.31%). The lifetime risk of RC was generally higher in more developed regions, including Northern Europe (2.76%), Western Europe (2.50%), Southern Europe (2.43%), and Eastern Europe (2.36%), as well as Australia/New Zealand where data are captured under high-HDI Western regions. In contrast, RC lifetime risk remained below 1.50% across most regions in Asia, Africa, Central and South America, and the Caribbean (Fig. 1). A broadly similar pattern was observed for CRC overall: CRC lifetime risk exceeded 7% in Northern, Western, and Southern Europe but remained below 3% across much of Africa, Central America, and Melanesia.

**Fig. 1:**
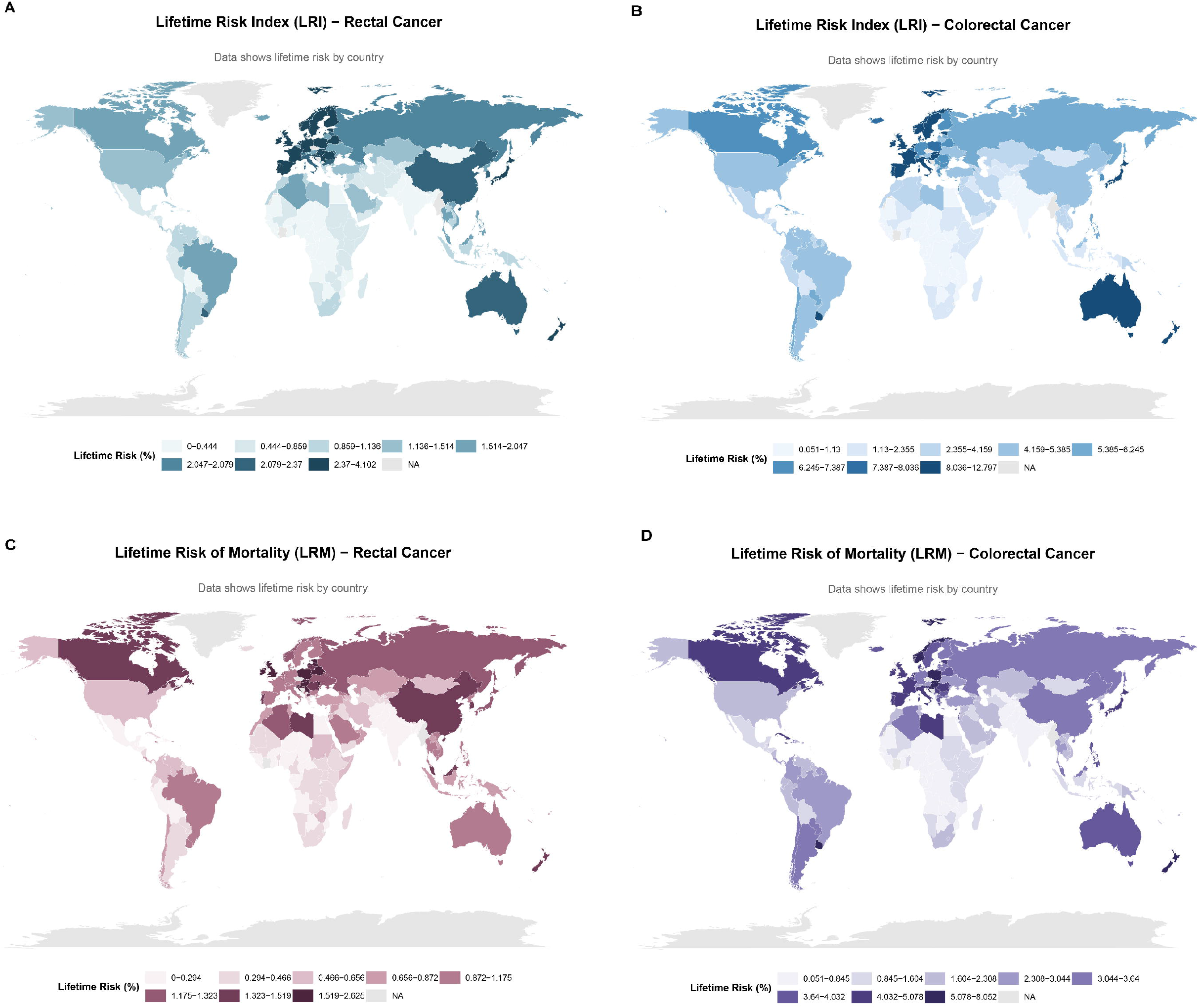
The global landscape of lifetime risks (%) of developing or dying from rectal and colorectal malignancies in 2022, both sexes. **Notes:** A. Developing (LRI) of RC; B. LRI of CRC. C. Dying from (LRM) of RC. D. LRM of CRC

However, the proportion of RC within the overall CRC burden (RC-to-CRC ratio) demonstrated a distinct geographic pattern that did not mirror the distribution of RC lifetime risk itself (Fig. 2). The RC-to-CRC ratio peaked in Central and South-Eastern Asia (41–43%), indicating a rectal-dominant CRC profile in these populations despite their moderate absolute RC risks. In contrast, regions such as the Caribbean and Central America exhibited the lowest ratios (<20%), reflecting a clear predominance of colon cancer even though their absolute RC risks were comparable to or higher than some African and Pacific regions. This divergence suggests that the relative anatomical distribution of CRC subsites is shaped by factors beyond those that determine absolute RC risk.

**Fig. 2:**
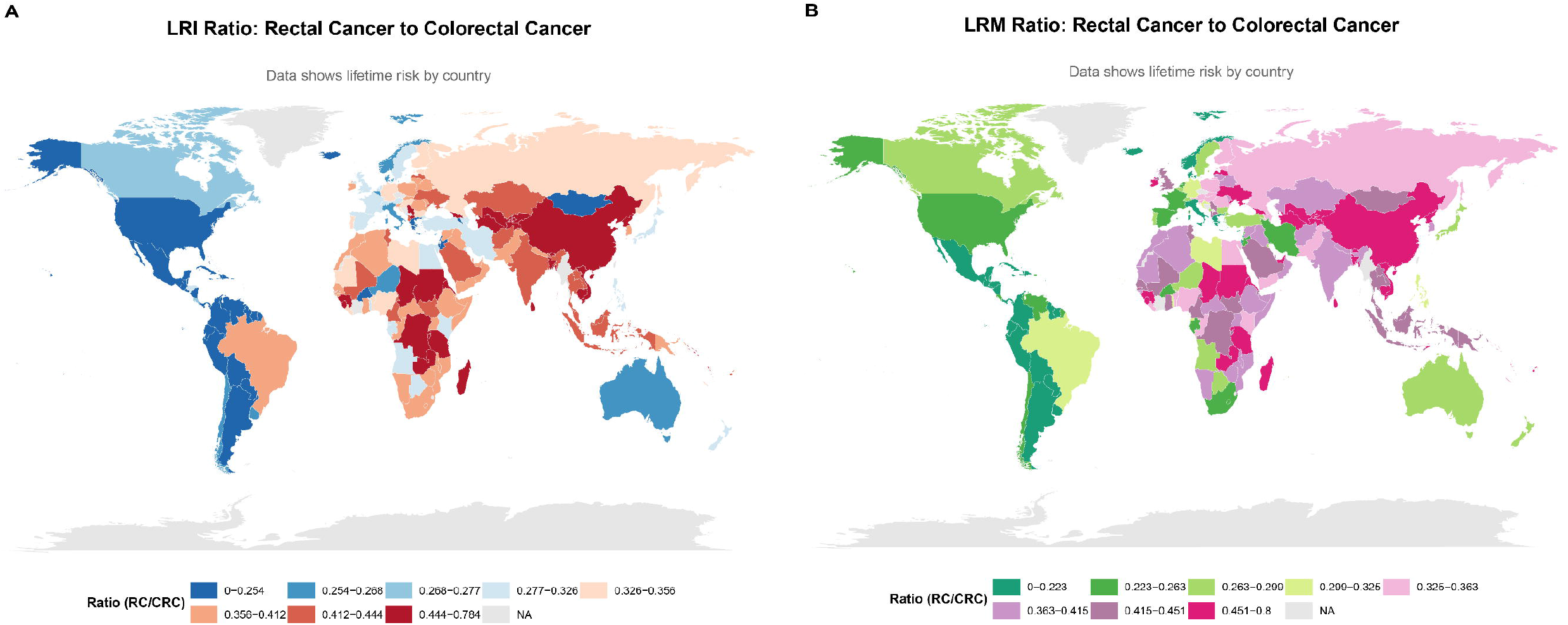
The Geographic pattern of the proportion of RC within the overall CRC burden. **Notes:** A. LRI ratio of RC/CRC. B. LRM ratio of RC/CRC

**Fig. 3.**
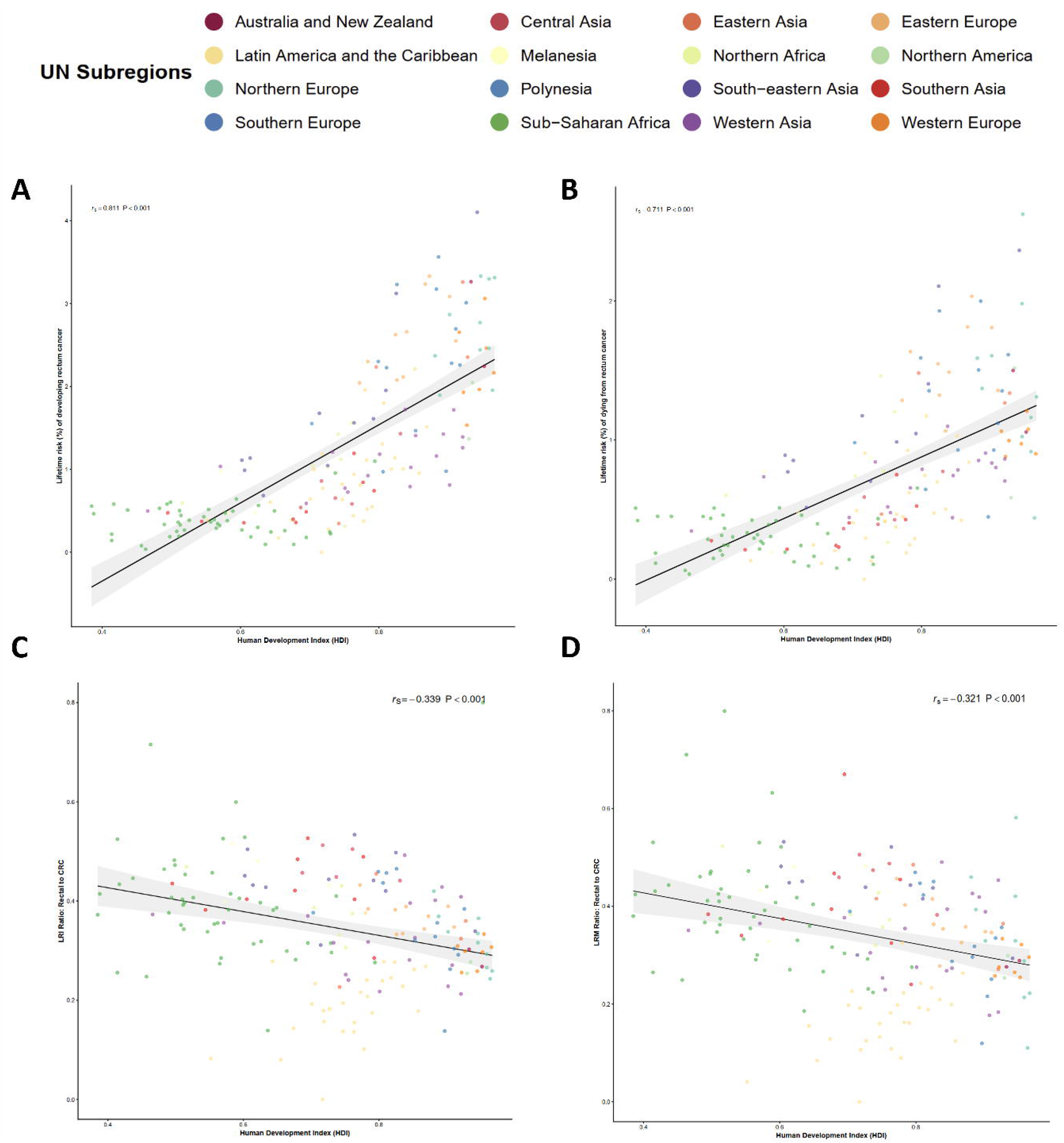
Association between lifetime risk indicators and the Human Development Index (HDI) across 185 countries. **Notes:** Panels show Spearman correlations between HDI and (A) lifetime risk of developing RC (LRI) in the total population; (B) lifetime risk of dying from RC (LRM) in the total population; (C) ratio of RC-to-CRC lifetime risk of developing cancer (LRI RC/CRC) in the total population; (D) ratio of RC-to-CRC lifetime risk of dying from cancer (LRM RC/CRC) in the total population.

Sex-stratified analyses followed the same geographical gradient, with higher RC and CRC lifetime risks in males across all regions (Supplementary Table S2-S3).

### Inequities in socioeconomic development and lifetime risk of colorectal malignancies

To further explore socioeconomic patterns, we analysed the association between HDI (as a continuous variable) and national lifetime risks across 185 countries. RC and CRC LRI and LRM were strongly positively correlated with HDI (all p<0.001; RC LRI *r*_*s*_=0.711; CRC LRI *r*_*s*_ =0.811), confirming a global socioeconomic gradient in absolute colorectal cancer burden (Figure 4A-B).

**Fig. 4:**
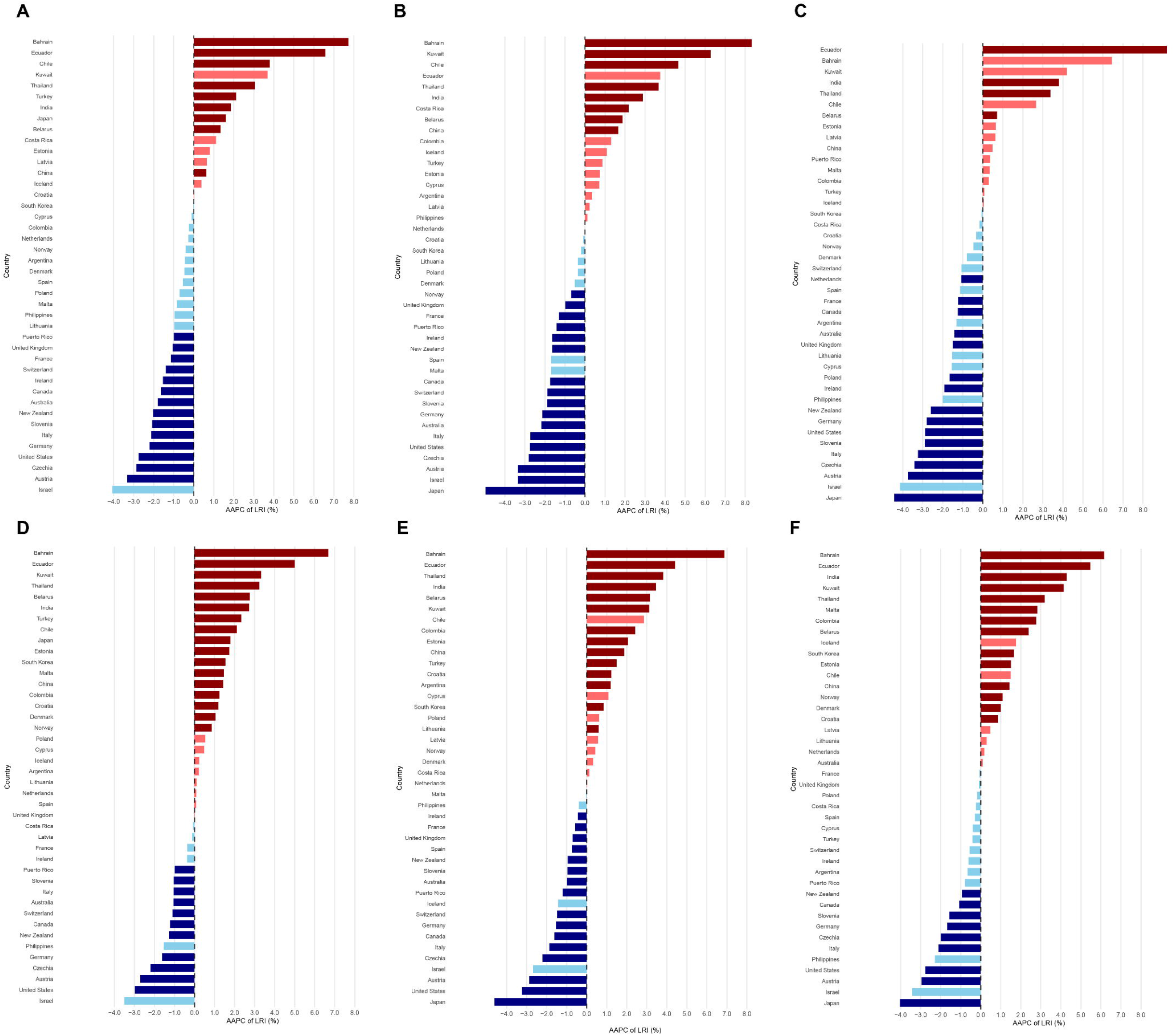
The average annual percent change (AAPC) for lifetime risks of developing or dying from colorectal malignancies from 2003 to 2017 by country, both sexes. Notes: A. Developing (LRI) rectal cancer of both sexes; B. Developing (LRI) rectal cancer of male; C. Developing (LRI) rectal cancer of female; D. Developing (LRI) colorectal cancer of both sexes; E. Developing (LRI) colorectal cancer of male; F. Developing (LRI) colorectal cancer of female. Dark red bars represent countries with a statistically significant increasing trend (AAPC > 0, p < 0.05). Light red bars represent countries with an increasing trend that is not statistically significant (AAPC > 0, p ≥ 0.05). Dark blue bars represent countries with a statistically significant decreasing trend (AAPC < 0, p < 0.05). Light blue bars represent the countries with a decreasing trend that is not statistically significant (AAPC < 0, p ≥ 0.05). AAPC: average annual percent change (%).

Conversely, the RC-to-CRC ratio was negatively associated with HDI, for both incidence and mortality (LRI ratio *r*_*s*_=−0.339, p<0.001; LRM ratio *r*_*s*_=−0.321, p<0.001), indicating that countries with lower socioeconomic development tend to have a more rectal-dominant CRC pattern (Figure 4C-D). This negative association persisted after sex stratification and was stronger among females (*r*_*s*_ =−0.342, p<0.001) than males (*r*_*s*_ =−0.237, p=0.002).

### LRI/LRM ratio

Analysis of the LRI/LRM ratio revealed broadly similar patterns for RC and CRC across HDI strata and geographic regions. The ratio increased with socioeconomic development for both cancers, with the highest values observed in very high HDI countries (RC 2.01; CRC 1.94) and the lowest in low HDI settings (RC 1.33; CRC 1.31). Regionally, high ratios for both cancers were concentrated in Northern America, Australia/New Zealand, and Europe—particularly Western and Southern Europe—whereas intermediate levels were observed in South America, Eastern Asia, Central America, the Caribbean, and Polynesia, and the lowest ratios were seen in Africa, Melanesia, and Micronesia (Supplementary Table S4).

However, greater divergence emerged at the national level. Substantial between-country heterogeneity and discordance were observed between RC and CRC, indicating that national performance in colorectal cancer overall did not consistently parallel rectal cancer–specific outcomes. The composition of countries with the highest RC ratios differed from that of CRC. Denmark and the Netherlands ranked highly for both cancers; however, Iceland had the highest RC ratio globally (4.44) but did not rank within the top ten for CRC. Haiti (2.92) and Suriname (2.62) were also among the five highest for RC despite comparatively moderate CRC ratios. Similarly, countries with the lowest RC ratios did not consistently correspond to those with the lowest CRC ratios. Mongolia (0.74) and Bahrain (0.97) had the lowest RC ratios, whereas Cape Verde showed low ratios for both cancers. Overall, the between-country range was wider for RC than for CRC, with RC ratios spanning from 0.74 in Mongolia to 4.44 in Iceland.

### Trend Analysis of LRI and LRM

In trend analysis, among the 42 countries with eligible consecutive surveillance data, significant heterogeneity in temporal trends was observed between RC and CRC. For RC LRI, 9 countries (21.4%) exhibited significant increases, with AAPCs ranging from 0.64% to 7.71%, while 14 countries (33.3%) showed significant declines. The most pronounced increase was observed in Bahrain (AAPC = 7.71%), whereas Austria demonstrated the largest decrease (AAPC = −3.32%). In contrast, increasing trends were more frequently observed for CRC LRI, with 17 countries (40.5%) showing significant rises (AAPCs 0.85%–6.68%), compared with 11 countries (26.2%) exhibiting significant declines. Bahrain again showed the most rapid increase (AAPC = 6.68%), while the United States experienced one of the most marked reductions (AAPC = −2.98%).

A similar pattern was observed for lifetime risk of mortality. For RC mortality, 10 countries (23.8%) exhibited significant increases (AAPCs 0.56%–10.55%), whereas 13 countries (31.0%) showed significant decreases. In contrast, CRC mortality increased in 16 countries (38.1%) and decreased in 10 countries (23.8%). Bahrain demonstrated the steepest increases in mortality trends, while the United States experienced the most pronounced declines. Overall, increasing trends were more frequently observed for CRC than for RC, whereas RC showed a greater proportion of declining trends across countries.

Sex-specific analyses revealed notable differences in LRI trends, particularly for RC. For RC LRI, significant increases were more frequently observed in males (8 countries) than in females (4 countries), whereas the number of countries with significant declines was similar between sexes (17 in males vs 15 in females). In contrast, LRI of CRC showed comparable numbers of increasing trends in both sexes (14 countries each), although significant decreases were more frequently observed in males (16 countries) than in females (9 countries). For lifetime risk of mortality, RC exhibited a more favorable pattern among females, with significant declines observed in 18 countries compared with 13 countries in males. Conversely, sex differences in colorectal cancer mortality trends were minimal, with similar numbers of countries showing increasing and decreasing trends in males and females (Fig. 4). LRM trends are similar to LRI trends (**Supplementary Figure 1**)

Regionally, increasing trends were concentrated in Eastern Asia and Latin America. Eastern Asia showed consistent increases for both CRC (mean AAPC 1.59%) and RC (mean AAPC 1.12%), driven by China, Japan, and the Republic of Korea. Latin America and the Caribbean demonstrated some of the fastest increases globally, including CRC (mean AAPC 2.78%) and particularly RC (mean AAPC 5.18%) in Chile and Ecuador. In contrast, Australia/New Zealand showed uniform declines for both CRC (mean AAPC −1.16%) and RC (mean AAPC −1.91%).

### Sensitivity analysis of lifetime risk versus 0–74 year cumulative risk

We observed strong linear correlations between lifetime risks and 0–74 year cumulative risks for both RC and CRC across 185 countries (r = 0.92–0.98, p < 0.001). In countries with low life expectancy (<75 years), lifetime risks were nearly identical to 0–74 year cumulative risks. However, in high-life-expectancy settings (>80 years), lifetime risks substantially exceeded 0–74 year cumulative risks, reflecting the additional contribution of cancer burden arising after age 75. The largest discrepancies were observed in Japan, Italy, and Singapore, where lifetime CRC risk was higher than the 0–74 year cumulative risk.

## Discussion

In this study, we performed a comprehensive global, regional and national assessment of the lifetime risks of RC and CRC using GLOBOCAN 2022 data. Several important patterns emerged. First, RC represents a substantial component of the global CRC burden, accounting for approximately 35% of lifetime CRC incidence and mortality worldwide. Second, marked geographical and socioeconomic heterogeneity was observed in both the absolute risks of RC and CRC and in the proportional distribution of CRC subsites. Third, although absolute lifetime risks increased with socioeconomic development, the proportional contribution of RC to total CRC showed an inverse socioeconomic pattern, with low socioeconomic development countries tending to exhibit a rectal cancer–dominant CRC profile. Finally, temporal trend analyses revealed considerable heterogeneity across countries, with increasing trends more frequently observed for CRC than for RC, whereas RC exhibited a greater proportion of declining trends across countries. Together, these findings suggest that rectal cancer constitutes not merely a subset of colorectal malignancy but an epidemiologically distinct component of the global colorectal cancer burden, and that socioeconomic development may increase the overall colorectal cancer burden while simultaneously reshaping its subsite spectrum.

Previous research has shown substantial variation in CRC incidence by anatomic subsite, although most available evidence comes from regional or single-country studies rather than global comparative analyses. Multiple population-based studies from high-income countries have reported that colon cancers—particularly proximal colon tumors—account for a growing share of the CRC burden in settings with established screening programs and ageing populations. For example, analyses from Australia, Canada, Denmark, New Zealand, and the United Kingdom indicate that approximately two-thirds of CRC cases arise in the colon, with proximal lesions increasing disproportionately in recent decades^[15]^. In contrast, several studies from low- and middle-income settings have reported a different subsite distribution. Hospital-based series from India^[16]^, parts of Sub-Saharan Africa^[17]^, and other transitioning regions show a higher proportion of distal and rectal cancers, with rectal tumours accounting for up to 50–60% of CRC in some populations. However, these studies relied on incidence rates and did not examine lifetime-risk patterns or quantify the proportional contribution of rectal disease across diverse life expectancy profiles. Our study extends this evidence by demonstrating that the RC-to-CRC ratio varies substantially across regions and, crucially, is negatively correlated with HDI, highlighting a discordance between absolute disease burden and subsite composition.

Absolute lifetime risks of both RC and CRC increased with HDI, reflecting well-established effects of population ageing, longer life expectancy, improved detection, and Westernized lifestyle exposures that increase cumulative cancer risk. However, the proportional contribution of RC within CRC did not increase in parallel with absolute risk and instead showed an inverse association with HDI, indicating that the determinants of total disease burden are not identical to those shaping its subsite distribution. One plausible explanation relates to differences in screening modality and penetration across regions. In settings with widespread colonoscopy-based screening, earlier detection and removal of colonic adenomas may disproportionately reduce colon cancer risk, thereby altering the relative balance between colon and rectal cancers. Conversely, in regions where screening is less widely implemented or relies more heavily on stool-based testing or limited endoscopic coverage, protection against proximal colonic neoplasia may be less consistent. Importantly, screening alone is unlikely to fully explain this pattern. The observed discordance more broadly supports the view that colon and rectal cancers are not merely adjacent anatomical manifestations of a single disease but may reflect partly distinct etiologic ecosystems shaped by differential screening exposure, diet, obesity, alcohol and tobacco use, environmental toxicants, microbiome composition, and molecular susceptibility. In this context, environmental and behavioral exposures may alter the anatomical distribution of tumors rather than simply increasing overall incidence, ultimately contributing to the complex inverse HDI gradient observed in subsite composition.

Importantly, the RC-to-CRC ratio displayed a geographic pattern that did not mirror absolute RC burden. Regions such as Central and South-Eastern Asia exhibited some of the highest RC-to-CRC ratios globally (approximately 41%–43%), despite not always having the highest absolute lifetime risks, suggesting that region-specific behavioral or environmental exposures may influence the anatomical distribution of CRC. Conversely, the Caribbean and parts of Central America demonstrated some of the lowest ratios (generally <20%), reflecting a colon-dominant CRC profile even in settings with moderate absolute risk levels. This geographic divergence illustrates that regions with similar overall CRC burdens may nonetheless display markedly different subsite distributions. The divergence between absolute risk and proportional subsite composition therefore suggests that the determinants of overall CRC incidence do not fully explain the anatomical distribution of disease, reinforcing the need for subsite-specific epidemiologic investigation and regionally tailored prevention strategies.

The LRI/LRM ratio, used here as a proxy indicator of survival in the absence of comprehensive population-based survival data, also revealed clear socioeconomic gradients. Both RC and CRC demonstrated higher ratios in more socioeconomically developed regions, consistent with improved early detection, greater treatment capacity and broader access to multimodality care in high-HDI countries. However, substantial between-country heterogeneity emerged at the national level, particularly for RC. The wider dispersion of RC ratios compared with CRC suggests that outcomes for rectal cancer may be more sensitive to differences in healthcare organization and treatment quality. This observation is biologically and clinically plausible. Rectal cancer management relies heavily on technically demanding procedures such as total mesorectal excision, accurate preoperative staging and coordinated multidisciplinary care, including neoadjuvant chemoradiotherapy. These components require specialized surgical expertise, radiotherapy infrastructure and centralized treatment systems that may vary widely across countries, even among those with similar overall healthcare capacity. In this context, rectal cancer outcomes may represent a more sensitive indicator of healthcare-system performance in colorectal oncology.

Temporal trend analysis further highlighted substantial heterogeneity across countries and regions. Increasing trends in lifetime CRC risk were observed more frequently than for RC, whereas RC showed a greater proportion of declining trends across countries. Regionally, increases were concentrated in Eastern Asia and parts of Latin America, with countries such as China, Japan, the Republic of Korea, Chile and Ecuador contributing to the rising burden. In contrast, several very-high HDI settings—including Australia and New Zealand—demonstrated consistent declines for both CRC and RC. These divergent trajectories likely reflect differing stages of epidemiologic transition. Rapidly developing economies may be experiencing increasing colorectal cancer burden driven by westernization of diet, rising obesity prevalence and changing patterns of alcohol and tobacco use. Meanwhile, long-standing high-income settings may be benefiting from screening implementation, improved risk-factor control and advances in treatment.

The findings of this study have several important implications for global cancer control. First, the discordance between absolute risk and proportional subsite burden suggests that rectal and colon cancers may be influenced by partially distinct etiologic pathways. Second, the substantial geographic variation in RC-to-CRC ratio indicates that prevention strategies may need to account for local subsite distribution patterns rather than relying solely on aggregated CRC statistics. Third, the observed divergence between RC and CRC outcomes at the national level suggests that improvements in overall colorectal cancer care may not uniformly translate into equivalent gains for rectal cancer. Strengthening multidisciplinary treatment capacity and ensuring access to specialized rectal cancer surgery may therefore represent important priorities for health systems.

The principal strength of this study lies in its comprehensive global estimation of lifetime risks of rectal and colorectal cancer across 185 countries using a standardized AMP framework integrating incidence, competing mortality, age-specific attenuation and temporal trends. This interpretation is further supported by our sensitivity analysis showing that lifetime risk diverged increasingly from 0–74-year cumulative risk in countries with longer life expectancy, underscoring the added value of lifetime-risk metrics in ageing populations. Nevertheless, several limitations should be acknowledged. Lifetime-risk estimates depend on the completeness and quality of underlying cancer registry data, and under-ascertainment persists in several low-resource regions. The AMP framework does not distinguish between proximal and distal colon cancers or incorporate molecular subtype information, limiting mechanistic interpretation. Trend analyses were restricted to countries with consecutive surveillance data, potentially biasing trend estimates toward higher-HDI settings. Finally, ecological associations between HDI and cancer burden cannot establish causality and likely reflect a complex interplay of demographic, environmental and healthcare-related factors.

In conclusion, by quantifying lifetime probabilities of developing and dying from rectal and colorectal cancers worldwide and characterizing their geographic patterns, socioeconomic gradients and temporal trends, this study highlights the substantial global burden of rectal cancer and underscores its epidemiologic distinctiveness within the broader colorectal cancer spectrum. These findings support the need for subsite-specific epidemiologic research, improved surveillance of rectal cancer outcomes and tailored prevention and treatment strategies across diverse population settings.

## Data Availability

The data used in this study are publicly available from GLOBOCAN 2022, the United Nations World Population Prospects, the United Nations Development Programme Human Development Index database, and the Cancer Incidence in Five Continents Plus (CI5plus) database. Derived data generated during the study are available from the corresponding author upon reasonable request.

https://gco.iarc.who.int

https://population.un.org/wpp/

https://hdr.undp.org/

https://ci5.iarc.fr/ci5plus/

---

**Table 1.**
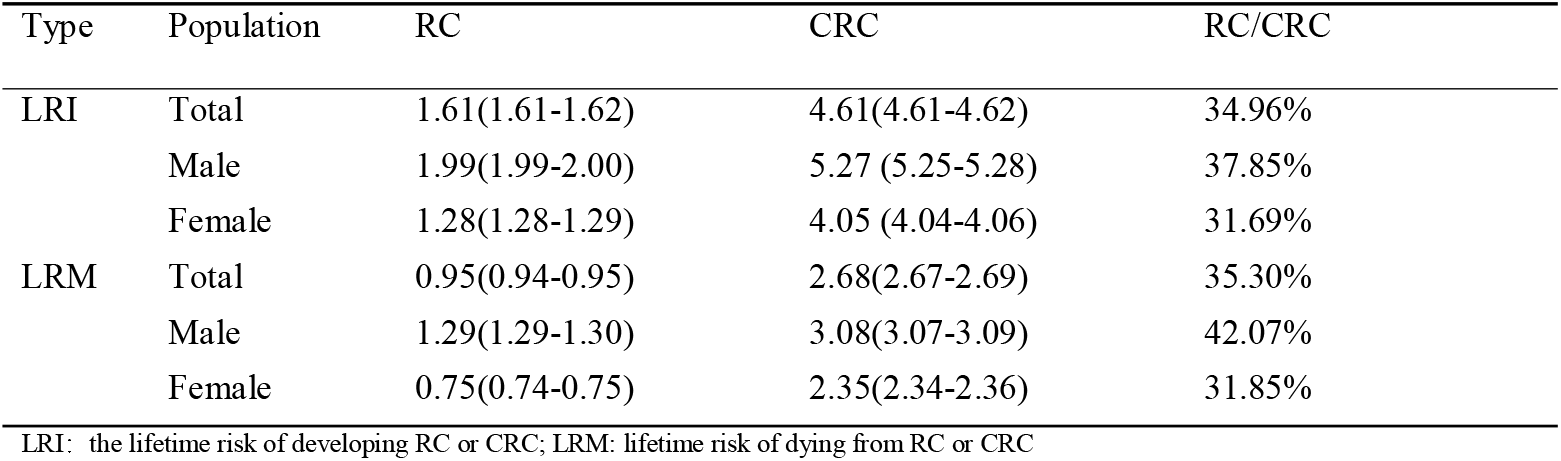
Lifetime risks (%) of developing and dying from colorectal cancers in 2022, by sex.

| Type | Population | RC | CRC | RC/CRC |
| --- | --- | --- | --- | --- |
| LRI | Total | 1.61(1.61-1.62) | 4.61(4.61-4.62) | 34.96% |
|  | Male | 1.99(1.99-2.00) | 5.27 (5.25-5.28) | 37.85% |
|  | Female | 1.28(1.28-1.29) | 4.05 (4.04-4.06) | 31.69% |
| LRM | Total | 0.95(0.94-0.95) | 2.68(2.67-2.69) | 35.30% |
|  | Male | 1.29(1.29-1.30) | 3.08(3.07-3.09) | 42.07% |
|  | Female | 0.75(0.74-0.75) | 2.35(2.34-2.36) | 31.85% |

LRI: the lifetime risk of developing RC or CRC; LRM: lifetime risk of dying from RC or CRC

