## Supplementary Material for "Distinct Global Patterns and Trends in Lifetime Risk of Rectal Cancer Within Colorectal Cancer: A Population-Based Analysis from GLOBOCAN 2022"

### Methods for calculating lifetime risk of developing or dying from cancer

The lifetime risks of developing and dying from cancer denote the probabilities of being diagnosed and dying from cancer. Generally, cumulative rate and cumulative risk are regarded as an approximation. Nevertheless, the two indicators are ideal for neglecting competing risks of death other than cancers, multiple primary cancers and life expectancy. The AMP method in the study corrects the above problems.

Here we provide a brief description of the methods of calculating lifetime risks of developing and dying from cancer as follows:

1. Cumulative rate

$$Cumulative rate=\sum_{i=0}^{A} W_{i}R_{i}$$

Where A is age-band, usually use 74 years as the upper limit for cumulative rate calculate; W_i_ is the width of the i^th^ age-band; R_i_ is the age-specific incidence rate or mortality rate in the i^th^ age-band.

2. Cumulative risk

$$Cumulative risk=1-e^{-cumulative rate}$$

3. The “adjusted for multiple primaries (AMP)” method^1,2^

The incidence data from the registry present the issues of multiple primary cancers being diagnosed in one person. To estimate the risks, we made the following assumptions: (1) one cannot die of cancer in a state where one never had cancer; (2) people who have never had cancer dying from causes other than cancer are identical to the general population; (3) the probability of developing cancer for those who have never had cancer before is comparable with the general population.

Mathematically, AMP method can be written as:

$$P=\int_{0}^{\infty} \lambda_{0c}S_{0}(a)da$$

Where P is the lifetime risk of developing cancer; λ_0c_ is the cancer incidence rate at age a; S_0_(a) is the probability of being alive and cancer-free at age a. In practice, we cannot directly estimate λ_0c_(a) and S_0_(a) from routine cancer registry data, and the data is aggregated in 5-year age groups (0-4, 5-9, …, 65-69, 70-84, and 85+ years), so we calculated with formula as follows:

$$P=\int_{0}^{\infty} \hat{\lambda}_{C}(a)\hat{S}_{0}^{*}(a)da=\sum_{i=1}^{f} \frac{R_{i}}{R_{i}+M_{i}-D_{i}}\hat{S}_{0}^{*}(a_{i})\times\left( 1-\exp(-\frac{W_{i}}{N_{i}}(R_{i}+M_{i}-D_{i})) \right)$$

Where for age group i:

Ri is the annual number of cancer cases (estimating the risk of developing cancer) or annual number of cancer deaths (estimating the risk of dying from cancer); M_i_ is the annual number of deaths (all-cause mortality); Di is the annual number of cancer deaths; Ni is the population; W_i_ is the width of the i^th^ age group, where the i^th^ interval is from a_i_ to a_i+1_, so W_i_=(a_i+1_-a_i_); $\hat{\lambda}_{c}$is the observed cancer incidence rate; $\hat{S}_{0}^{*}(a_{i})$ is the probability of being alive and cancer free at age a_i_.

$$\hat{S}_{0}^{*}(a_{i})=exp(-\sum_{j=1}^{i-1} \frac{R_{j}+(M_{j}-D_{j})}{N_{j}})$$

Generally, the final age band of cancer incidence and mortality is usually 85 years old or above. For the final age band (e.g. 85+), the integral for the final age band as follows:

$$\frac{R_{f}}{R_{f}+M_{f}-D_{f}}S_{0}(a_{f})$$

The 95% confidence interval (CI) of lifetime risk was estimated based on the poisson method. The derivation of variance as follows:

let: $\mu_{c}=\frac{R_{i}}{R_{i}+M_{i}-D_{i}},\hat{S}_{0}^{*}=\hat{S}_{0}^{*}\left( a_{i} \right),S_{x}=\left( 1-\exp(-\frac{W_{i}}{N_{i}}(R_{i}+M_{i}-D_{i})) \right)$

then

$$Var\left( S \right)=\sum_{i=1}^{f} Var\left( \mu_{c}\hat{S}_{0}^{*}S_{x} \right)=\sum_{i=1}^{f} \left[ E\left( \mu_{c}^{2}\left( \hat{S}_{0}^{*} \right)^{2}S_{x}^{2} \right)-{E\left( \mu_{c}\hat{S}_{0}^{*}S_{x} \right)}^{2} \right] (1)$$

$$E\left( \mu_{c}^{2}\left( \hat{S}_{0}^{*} \right)^{2}S_{x}^{2} \right)=E(\mu_{c}^{2})E\left( \hat{S}_{0}^{*} \right)^{2}E\left( S_{x}^{2} \right)=\left( [var(\hat{S}_{0}^{*})+{(E\hat{S}_{0}^{*})}^{2}][Var\left( S_{x} \right)+{E(S_{x})}^{2}]\} \right)$$

$$Var\left( S \right)=\sum_{i=1}^{f} \{[Var (\mu_{c})+\left( E\mu_{c} \right)^{2}][Var\left( \hat{S}_{0}^{*} \right)+\left( E\hat{S}_{0}^{*} \right)^{2}][Var\left( S_{x} \right)+\left( ES_{x} \right)^{2}-\left( E\mu_{c} \right)^{2}\left( E\hat{S}_{0}^{*} \right)^{2}\left( ES_{x} \right)^{2}]\}$$

let: $p_{ci}=\frac{R_{i}}{R_{i}+M_{i}-D_{i}}$, then

$$E\mu_{c}=p_{ci} (2)$$

$$Var\left( \mu_{c} \right)=\frac{p_{ci}}{R_{i}+M_{i}-D_{i}} (3)$$

$\log\left( \hat{S}_{0}^{*} \right)=-\sum_{j=1}^{i-1} \frac{R_{j}+M_{j}-D_{j}}{N_{j}}$,

let $p_{oj}=\frac{R_{j}+M_{j}-D_{j}}{N_{j}}$, then

$$Var\left( log\hat{S}_{0}^{*} \right)=\sum_{j=1}^{i-1} Var\left( \frac{R_{j}+M_{j}-D_{j}}{N_{j}} \right)=\sum_{j=1}^{i-1} \frac{p_{oj}}{N_{j}}$$

According to delta method, if $\hat{\theta}\sim N\left( \theta,\sigma^{2} \right)$, then $f\left( \hat{\theta} \right)\sim N (f\left( \theta\right),{[f^{'}\left( \theta\right)]}^{2}\sigma^{2})$. Let $\hat{S}_{0}^{*}=e^{\hat{\theta}}$, then

$$E\left( \hat{S}_{0}^{*} \right)=\exp\left( -\sum_{j=1}^{i-1} p_{oj} \right) (4)$$

$$Var\left( \hat{S}_{0}^{*} \right)={(e^{\hat{\theta}})}^{2}Var\left( log\hat{S}_{0}^{*} \right)=\left( \hat{S}_{0}^{*} \right)^{2}Var\left( log\hat{S}_{0}^{*} \right)=\left( \hat{S}_{0}^{*} \right)^{2}\times\sum_{j=1}^{i-1} \frac{p_{oj}}{N_{j}} (5)$$

$S_{x}=\left( 1-\exp\left( -\frac{w_{i}}{N_{i}}\left( R_{i}+M_{i}-D_{i} \right) \right) \right)$,

let $S_{x}^{'}=\exp\left( -\frac{w_{i}}{N_{i}}\left( R_{i}+M_{i}-D_{i} \right) \right)$, $\log\left( S_{x}^{'} \right)=-\frac{w_{i}}{N_{i}}\left( R_{i}+M_{i}-D_{i} \right)$, and $p_{xi}=\frac{R_{i}}{R_{i}+M_{i}-D_{i}}$, then

$$Var(\log S_{x}^{'})={w_{i}}^{2}\frac{p_{xi}}{N_{i}}$$

let $S_{x}=1-e^{\hat{\theta}}$, according to delta method, then:

$$E\left( S_{x} \right)=\left( 1-\exp\left( -\frac{w_{i}}{N_{i}}\left( R_{i}+M_{i}-D_{i} \right) \right) \right) (6)$$

$$Var\left( S_{x} \right)={(e^{\hat{\theta}})}^{2}\times Var\left( e^{\hat{\theta}} \right)={(S_{x}^{'})}^{2}\times{w_{i}}^{2}\frac{p_{xi}}{N_{i}} (7)$$

### SP Table 1. Cancer classification for colorectal malignancies used in lifetime risk analysis

| **ICD-10** | **Sites** | **Short title used in the paper** |
| --- | --- | --- |
| **C18-C21** | Colorectum | CRC |
| **C19-C20** | Rectum | RC |

### SP Table 2. Lifetime risks (%) of developing (LRI) colorectal malignancies in 2022, by sex, HDI level and region

| **Type** | **Population** | **Rectum** | **Colorectum** | **RC/CRC** |
| --- | --- | --- | --- | --- |
| **Both** | **HDI level** |  |  |  |
|  | Very high HDI | 2.27 (2.26-2.28) | 7.38 (7.36-7.40) | 30.73% |
|  | High HDI | 1.73 (1.72-1.74) | 4.08 (4.07-4.09) | 42.48% |
|  | Medium HDI | 0.54 (0.53-0.55) | 1.39 (1.38-1.40) | 38.78% |
|  | Low HDI | 0.58 (0.57-0.60) | 1.54 (1.51-1.56) | 37.95% |
|  | **Region** |  |  |  |
|  | Eastern Africa | 0.53 (0.51-0.55) | 1.33 (1.30-1.36) | 39.68% |
|  | Middle Africa | 0.35 (0.33-0.37) | 0.82 (0.78-0.85) | 42.82% |
|  | Northern Africa | 0.82 (0.80-0.85) | 2.20 (2.16-2.25) | 37.35% |
|  | Southern Africa | 0.91 (0.86-0.95) | 2.22 (2.16-2.29) | 40.72% |
|  | Western Africa | 0.34 (0.32-0.35) | 0.95 (0.92-0.97) | 35.38% |
|  | Eastern Asia | 2.32 (2.31-2.33) | 5.65 (5.64-5.67) | 41.06% |
|  | South-Eastern Asia | 1.37 (1.35-1.39) | 3.19 (3.17-3.22) | 42.90% |
|  | Western Asia | 1.17 (1.15-1.20) | 3.70 (3.65-3.75) | 31.64% |
|  | Eastern Europe | 2.36 (2.34-2.38) | 6.53 (6.49-6.56) | 36.10% |
|  | Northern Europe | 2.76 (2.72-2.80) | 8.87 (8.80-8.94) | 31.10% |
|  | Southern Europe | 2.43 (2.40-2.45) | 8.39 (8.34-8.44) | 28.95% |
|  | Western Europe | 2.50 (2.48-2.52) | 7.94 (7.89-7.98) | 31.49% |
|  | Caribbean | 0.86 (0.82-0.90) | 5.18 (5.07-5.30) | 16.62% |
|  | Central America | 0.53 (0.51-0.55) | 2.62 (2.57-2.67) | 20.40% |
|  | South America | 1.47 (1.45-1.49) | 4.89 (4.85-4.92) | 30.01% |
|  | Northern America | 1.44 (1.43-1.46) | 5.61 (5.58-5.64) | 25.73% |
|  | Australia/New Zealand | 2.41 (2.34-2.48) | 8.76 (8.63-8.90) | 27.48% |
|  | Melanesia | 0.98 (0.82-1.13) | 2.44 (2.20-2.69) | 39.97% |
|  | Micronesia | 1.18 (0.71-1.66) | 4.50 (3.14-5.87) | 26.29% |
|  | Polynesia | 1.53 (0.96-2.09) | 3.63 (2.78-4.48) | 42.03% |
| **Male** | **HDI level** |  |  |  |
|  | Very high HDI | 2.83 (2.81-2.84) | 8.36 (8.33-8.38) | 33.83% |
|  | High HDI | 2.18 (2.17-2.19) | 4.85 (4.84-4.87) | 44.94% |
|  | Medium HDI | 0.63 (0.62-0.64) | 1.60 (1.58-1.62) | 39.18% |
|  | Low HDI | 0.63 (0.61-0.66) | 1.70 (1.67-1.74) | 37.27% |
|  | **Region** |  |  |  |
|  | Eastern Africa | 0.59 (0.56-0.62) | 1.49 (1.44-1.53) | 39.77% |
|  | Middle Africa | 0.37 (0.33-0.40) | 0.83 (0.78-0.88) | 44.02% |
|  | Northern Africa | 0.89 (0.84-0.93) | 2.41 (2.34-2.48) | 36.70% |
|  | Southern Africa | 1.12 (1.05-1.20) | 2.78 (2.66-2.90) | 40.42% |
|  | Western Africa | 0.36 (0.34-0.38) | 1.05 (1.02-1.09) | 34.27% |
|  | Eastern Asia | 2.94 (2.92-2.95) | 6.61 (6.58-6.63) | 44.44% |
|  | South-Eastern Asia | 1.71 (1.68-1.74) | 3.85 (3.80-3.89) | 44.45% |
|  | Western Asia | 1.37 (1.33-1.41) | 3.93 (3.86-4.00) | 34.91% |
|  | Eastern Europe | 3.21 (3.17-3.25) | 8.10 (8.04-8.17) | 39.59% |
|  | Northern Europe | 3.47 (3.41-3.53) | 10.03 (9.92-10.14) | 34.56% |
|  | Southern Europe | 3.13 (3.09-3.18) | 10.14 (10.06-10.22) | 30.92% |
|  | Western Europe | 2.92 (2.89-2.96) | 8.84 (8.77-8.91) | 33.06% |
|  | Caribbean | 0.90 (0.83-0.96) | 4.95 (4.78-5.11) | 18.13% |
|  | Central America | 0.67 (0.63-0.70) | 2.80 (2.72-2.87) | 23.84% |
|  | South America | 1.67 (1.64-1.70) | 5.13 (5.07-5.18) | 32.59% |
|  | Northern America | 1.75 (1.73-1.78) | 5.97 (5.92-6.01) | 29.39% |
|  | Australia/New Zealand | 2.96 (2.85-3.06) | 9.30 (9.11-9.50) | 31.79% |
|  | Melanesia | 1.06 (0.83-1.29) | 2.88 (2.50-3.27) | 36.78% |
|  | Micronesia | 0.95 (0.41-1.49) | 4.40 (2.59-6.22) | 21.59% |
|  | Polynesia | 2.03 (1.07-3.00) | 4.54 (3.13-5.95) | 44.80% |
| **Female** | **HDI level** |  |  |  |
|  | Very high HDI | 1.79 (1.78-1.80) | 6.55 (6.53-6.57) | 27.29% |
|  | High HDI | 1.34 (1.33-1.35) | 3.41 (3.39-3.42) | 39.41% |
|  | Medium HDI | 0.46 (0.45-0.47) | 1.20 (1.19-1.22) | 38.20% |
|  | Low HDI | 0.54 (0.52-0.56) | 1.39 (1.36-1.42) | 38.66% |
|  | **Region** |  |  |  |
|  | Eastern Africa | 0.48 (0.46-0.50) | 1.21 (1.18-1.25) | 39.60% |
|  | Middle Africa | 0.34 (0.31-0.36) | 0.80 (0.75-0.85) | 41.89% |
|  | Northern Africa | 0.77 (0.73-0.80) | 2.03 (1.97-2.09) | 37.90% |
|  | Southern Africa | 0.76 (0.71-0.80) | 1.86 (1.78-1.94) | 40.67% |
|  | Western Africa | 0.31 (0.29-0.33) | 0.85 (0.82-0.88) | 36.57% |
|  | Eastern Asia | 1.76 (1.75-1.77) | 4.78 (4.76-4.80) | 36.90% |
|  | South-Eastern Asia | 1.11 (1.09-1.13) | 2.69 (2.65-2.72) | 41.30% |
|  | Western Asia | 0.99 (0.96-1.02) | 3.47 (3.41-3.54) | 28.52% |
|  | Eastern Europe | 1.80 (1.78-1.82) | 5.55 (5.51-5.59) | 32.41% |
|  | Northern Europe | 2.14 (2.10-2.19) | 7.88 (7.79-7.96) | 27.20% |
|  | Southern Europe | 1.82 (1.78-1.85) | 6.89 (6.83-6.96) | 26.33% |
|  | Western Europe | 2.10 (2.07-2.13) | 7.11 (7.06-7.17) | 29.47% |
|  | Caribbean | 0.83 (0.77-0.89) | 5.37 (5.21-5.53) | 15.47% |
|  | Central America | 0.42 (0.40-0.45) | 2.47 (2.40-2.53) | 17.15% |
|  | South America | 1.29 (1.27-1.32) | 4.68 (4.63-4.73) | 27.63% |
|  | Northern America | 1.16 (1.14-1.17) | 5.27 (5.23-5.31) | 21.95% |
|  | Australia/New Zealand | 1.89 (1.81-1.97) | 8.24 (8.05-8.42) | 22.96% |
|  | Melanesia | 0.89 (0.67-1.10) | 1.97 (1.67-2.27) | 44.92% |
|  | Micronesia | 1.36 (0.64-2.08) | 4.50 (2.54-6.47) | 30.25% |
|  | Polynesia | 1.06 (0.41-1.72) | 2.80 (1.77-3.83) | 37.99% |

HDI: Human Development Index.

### SP Table S3. Lifetime risks (%) of dying from (LRM) colorectal malignancies in 2022, by sex, HDI level and region

| **Type** | **Population** | **Rectum** | **Colorectum** | **RC/CRC** |
| --- | --- | --- | --- | --- |
| **Both** | **HDI level** |  |  |  |
|  | Very high HDI | 1.13 (1.12-1.13) | 3.80 (3.78-3.81) | 29.67% |
|  | High HDI | 1.12 (1.11-1.13) | 2.56 (2.55-2.57) | 43.78% |
|  | Medium HDI | 0.35 (0.34-0.35) | 0.92 (0.92-0.93) | 37.68% |
|  | Low HDI | 0.44 (0.43-0.45) | 1.17 (1.15-1.19) | 37.43% |
|  | **Region** |  |  |  |
|  | Eastern Africa | 0.42 (0.40-0.44) | 1.05 (1.02-1.08) | 40.05% |
|  | Middle Africa | 0.28 (0.26-0.30) | 0.67 (0.64-0.71) | 41.65% |
|  | Northern Africa | 0.66 (0.63-0.69) | 1.72 (1.67-1.76) | 38.43% |
|  | Southern Africa | 0.42 (0.39-0.45) | 1.85 (1.78-1.92) | 22.83% |
|  | Western Africa | 0.27 (0.26-0.28) | 0.75 (0.73-0.77) | 35.97% |
|  | Eastern Asia | 1.39 (1.38-1.40) | 3.28 (3.27-3.30) | 42.39% |
|  | South-Eastern Asia | 0.92 (0.91-0.94) | 2.18 (2.15-2.20) | 42.42% |
|  | Western Asia | 0.72 (0.70-0.74) | 2.47 (2.43-2.51) | 29.07% |
|  | Eastern Europe | 1.43 (1.42-1.45) | 3.91 (3.88-3.94) | 36.63% |
|  | Northern Europe | 1.73 (1.70-1.76) | 4.49 (4.44-4.55) | 38.48% |
|  | Southern Europe | 1.08 (1.06-1.10) | 4.35 (4.31-4.38) | 24.91% |
|  | Western Europe | 1.07 (1.05-1.08) | 3.69 (3.66-3.72) | 28.89% |
|  | Caribbean | 0.46 (0.43-0.50) | 3.52 (3.42-3.62) | 13.19% |
|  | Central America | 0.27 (0.25-0.28) | 1.61 (1.57-1.65) | 16.55% |
|  | South America | 0.71 (0.70-0.73) | 2.79 (2.76-2.82) | 25.51% |
|  | Northern America | 0.69 (0.68-0.70) | 2.62 (2.60-2.65) | 26.34% |
|  | Australia/New Zealand | 1.13 (1.08-1.18) | 3.95 (3.85-4.04) | 28.62% |
|  | Melanesia | 0.71 (0.57-0.86) | 1.68 (1.46-1.90) | 42.44% |
|  | Micronesia | 0.61 (0.26-0.97) | 3.43 (2.12-4.74) | 17.89% |
|  | Polynesia | 0.86 (0.49-1.23) | 1.88 (1.34-2.41) | 45.66% |
| **Male** | **HDI level** |  |  |  |
|  | Very high HDI | 1.43 (1.41-1.44) | 4.34 (4.32-4.36) | 32.87% |
|  | High HDI | 1.44 (1.42-1.45) | 3.06 (3.05-3.08) | 46.85% |
|  | Medium HDI | 0.40 (0.39-0.41) | 1.05 (1.04-1.07) | 37.74% |
|  | Low HDI | 0.46 (0.45-0.48) | 1.28 (1.25-1.31) | 36.26% |
|  | **Region** |  |  |  |
|  | Eastern Africa | 0.46 (0.43-0.49) | 1.16 (1.12-1.20) | 39.67% |
|  | Middle Africa | 0.29 (0.26-0.32) | 0.68 (0.63-0.73) | 42.58% |
|  | Northern Africa | 0.70 (0.66-0.74) | 1.90 (1.83-1.97) | 36.86% |
|  | Southern Africa | 0.48 (0.43-0.54) | 2.09 (1.97-2.21) | 23.09% |
|  | Western Africa | 0.29 (0.27-0.31) | 0.83 (0.80-0.87) | 34.29% |
|  | Eastern Asia | 1.77 (1.76-1.79) | 3.80 (3.78-3.83) | 46.55% |
|  | South-Eastern Asia | 1.16 (1.13-1.19) | 2.62 (2.58-2.66) | 44.24% |
|  | Western Asia | 0.80 (0.77-0.84) | 2.67 (2.61-2.74) | 29.97% |
|  | Eastern Europe | 2.00 (1.97-2.04) | 5.07 (5.01-5.12) | 39.54% |
|  | Northern Europe | 2.17 (2.12-2.23) | 5.07 (4.99-5.15) | 42.86% |
|  | Southern Europe | 1.41 (1.38-1.44) | 5.36 (5.30-5.43) | 26.28% |
|  | Western Europe | 1.36 (1.33-1.39) | 4.28 (4.23-4.33) | 31.78% |
|  | Caribbean | 0.49 (0.44-0.55) | 3.38 (3.24-3.53) | 14.58% |
|  | Central America | 0.33 (0.30-0.35) | 1.71 (1.65-1.77) | 19.02% |
|  | South America | 0.80 (0.78-0.83) | 2.94 (2.90-2.98) | 27.36% |
|  | Northern America | 0.86 (0.84-0.88) | 2.84 (2.80-2.87) | 30.31% |
|  | Australia/New Zealand | 1.39 (1.31-1.47) | 4.23 (4.08-4.38) | 32.92% |
|  | Melanesia | 0.80 (0.58-1.02) | 2.07 (1.72-2.43) | 38.58% |
|  | Micronesia | 0.40 (0.13-0.67) | 3.28 (1.56-5.01) | 12.29% |
|  | Polynesia | 1.21 (0.56-1.87) | 2.46 (1.54-3.38) | 49.24% |
| **Female** | **HDI level** |  |  |  |
|  | Very high HDI | 0.88 (0.88-0.89) | 3.35 (3.34-3.37) | 26.32% |
|  | High HDI | 0.86 (0.85-0.87) | 2.14 (2.12-2.15) | 40.10% |
|  | Medium HDI | 0.30 (0.30-0.31) | 0.81 (0.80-0.82) | 37.51% |
|  | Low HDI | 0.42 (0.40-0.43) | 1.08 (1.05-1.10) | 38.60% |
|  | **Region** |  |  |  |
|  | Eastern Africa | 0.38 (0.36-0.41) | 0.96 (0.93-1.00) | 39.92% |
|  | Middle Africa | 0.27 (0.24-0.30) | 0.67 (0.62-0.71) | 40.55% |
|  | Northern Africa | 0.63 (0.59-0.66) | 1.58 (1.52-1.64) | 39.74% |
|  | Southern Africa | 0.38 (0.34-0.42) | 1.70 (1.62-1.79) | 22.35% |
|  | Western Africa | 0.25 (0.24-0.27) | 0.68 (0.65-0.71) | 37.10% |
|  | Eastern Asia | 1.06 (1.05-1.07) | 2.82 (2.81-2.84) | 37.45% |
|  | South-Eastern Asia | 0.75 (0.73-0.77) | 1.85 (1.82-1.88) | 40.41% |
|  | Western Asia | 0.64 (0.61-0.67) | 2.30 (2.24-2.35) | 27.97% |
|  | Eastern Europe | 1.09 (1.07-1.11) | 3.24 (3.21-3.27) | 33.58% |
|  | Northern Europe | 1.36 (1.32-1.40) | 4.02 (3.95-4.08) | 33.91% |
|  | Southern Europe | 0.81 (0.79-0.84) | 3.52 (3.48-3.57) | 23.10% |
|  | Western Europe | 0.82 (0.80-0.84) | 3.20 (3.16-3.23) | 25.73% |
|  | Caribbean | 0.44 (0.39-0.48) | 3.62 (3.48-3.76) | 12.09% |
|  | Central America | 0.21 (0.20-0.23) | 1.52 (1.47-1.58) | 14.10% |
|  | South America | 0.63 (0.61-0.65) | 2.67 (2.63-2.70) | 23.74% |
|  | Northern America | 0.54 (0.53-0.56) | 2.42 (2.39-2.45) | 22.36% |
|  | Australia/New Zealand | 0.88 (0.82-0.95) | 3.67 (3.54-3.80) | 24.09% |
|  | Melanesia | 0.62 (0.43-0.81) | 1.27 (1.01-1.52) | 48.84% |
|  | Micronesia | 0.77 (0.18-1.35) | 3.48 (1.59-5.38) | 21.97% |
|  | Polynesia | 0.53 (0.13-0.92) | 1.35 (0.74-1.95) | 39.05% |

HDI: Human Development Index.

### SP Table 4. The LRI/LRM ratio for colorectal cancers in 2022, by sex, HDI level and region

| **Type** | **Population** | **Rectum** | **Colorectum** |
| --- | --- | --- | --- |
| **Both** | **HDI level** |  |  |
|  | Very high HDI | 2.01 | 1.94 |
|  | High HDI | 1.55 | 1.59 |
|  | Medium HDI | 1.55 | 1.51 |
|  | Low HDI | 1.33 | 1.31 |
|  | **Region** |  |  |
|  | Eastern Africa | 1.26 | 1.27 |
|  | Middle Africa | 1.25 | 1.21 |
|  | Northern Africa | 1.24 | 1.28 |
|  | Southern Africa | 2.14 | 1.20 |
|  | Western Africa | 1.24 | 1.26 |
|  | Eastern Asia | 1.67 | 1.72 |
|  | South-Eastern Asia | 1.48 | 1.47 |
|  | Western Asia | 1.63 | 1.50 |
|  | Eastern Europe | 1.65 | 1.67 |
|  | Northern Europe | 1.59 | 1.97 |
|  | Southern Europe | 2.24 | 1.93 |
|  | Western Europe | 2.34 | 2.15 |
|  | Caribbean | 1.86 | 1.47 |
|  | Central America | 2.00 | 1.62 |
|  | South America | 2.06 | 1.75 |
|  | Northern America | 2.09 | 2.14 |
|  | Australia/New Zealand | 2.13 | 2.22 |
|  | Melanesia | 1.37 | 1.45 |
|  | Micronesia | 1.93 | 1.31 |
|  | Polynesia | 1.78 | 1.94 |
| **Male** | **HDI level** |  |  |
|  | Very high HDI | 1.98 | 1.93 |
|  | High HDI | 1.52 | 1.58 |
|  | Medium HDI | 1.58 | 1.52 |
|  | Low HDI | 1.37 | 1.33 |
|  | **Region** |  |  |
|  | Eastern Africa | 1.28 | 1.28 |
|  | Middle Africa | 1.26 | 1.22 |
|  | Northern Africa | 1.27 | 1.27 |
|  | Southern Africa | 2.33 | 1.33 |
|  | Western Africa | 1.26 | 1.26 |
|  | Eastern Asia | 1.66 | 1.74 |
|  | South-Eastern Asia | 1.47 | 1.47 |
|  | Western Asia | 1.71 | 1.47 |
|  | Eastern Europe | 1.60 | 1.60 |
|  | Northern Europe | 1.60 | 1.98 |
|  | Southern Europe | 2.22 | 1.89 |
|  | Western Europe | 2.15 | 2.07 |
|  | Caribbean | 1.82 | 1.46 |
|  | Central America | 2.05 | 1.63 |
|  | South America | 2.08 | 1.74 |
|  | Northern America | 2.04 | 2.10 |
|  | Australia/New Zealand | 2.12 | 2.20 |
|  | Melanesia | 1.33 | 1.39 |
|  | Micronesia | 2.36 | 1.34 |
|  | Polynesia | 1.68 | 1.84 |
| **Female** | **HDI level** |  |  |
|  | Very high HDI | 2.02 | 1.95 |
|  | High HDI | 1.57 | 1.59 |
|  | Medium HDI | 1.52 | 1.49 |
|  | Low HDI | 1.29 | 1.29 |
|  | **Region** |  |  |
|  | Eastern Africa | 1.25 | 1.26 |
|  | Middle Africa | 1.24 | 1.20 |
|  | Northern Africa | 1.23 | 1.29 |
|  | Southern Africa | 1.99 | 1.09 |
|  | Western Africa | 1.23 | 1.25 |
|  | Eastern Asia | 1.67 | 1.69 |
|  | South-Eastern Asia | 1.48 | 1.45 |
|  | Western Asia | 1.54 | 1.51 |
|  | Eastern Europe | 1.65 | 1.71 |
|  | Northern Europe | 1.57 | 1.96 |
|  | Southern Europe | 2.23 | 1.96 |
|  | Western Europe | 2.55 | 2.23 |
|  | Caribbean | 1.90 | 1.48 |
|  | Central America | 1.97 | 1.62 |
|  | South America | 2.04 | 1.76 |
|  | Northern America | 2.14 | 2.18 |
|  | Australia/New Zealand | 2.14 | 2.25 |
|  | Melanesia | 1.43 | 1.56 |
|  | Micronesia | 1.78 | 1.29 |
|  | Polynesia | 2.02 | 2.08 |

HDI: Human Development Index.

### SP Figure 1. The average annual percent change (AAPC) for lifetime risks of dying from colorectal malignancies from 2003 to 2017 by country, both sexes.

The average annual percent change (AAPC) for lifetime risks of developing or dying from colorectal malignancies from 2003 to 2017 by country, both sexes. Notes: A. LRM of rectal cancer of both sexes; B. LRM of rectal cancer of male; C. LRM of female; D. LRM colorectal cancer of both sexes; E. LRM of colorectal cancer of male; F. LRM of colorectal cancer of female. Dark red bars represent countries with a statistically significant increasing trend (AAPC > 0, p < 0.05). Light red bars represent countries with an increasing trend that is not statistically significant (AAPC > 0, p ≥ 0.05). Dark blue bars represent countries with a statistically significant decreasing trend (AAPC < 0, p < 0.05). Light blue bars represent the countries with a decreasing trend that is not statistically significant (AAPC < 0, p ≥ 0.05). AAPC: average annual percent change (%).

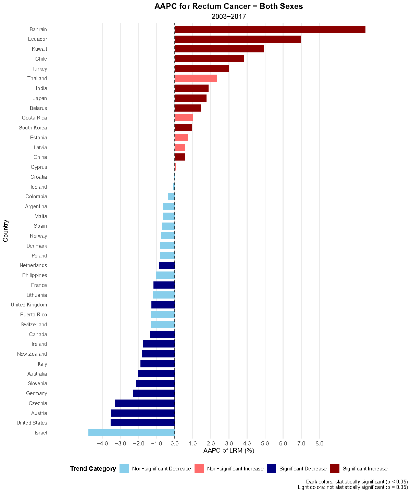

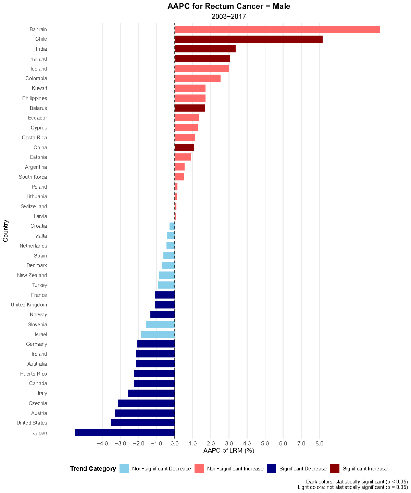

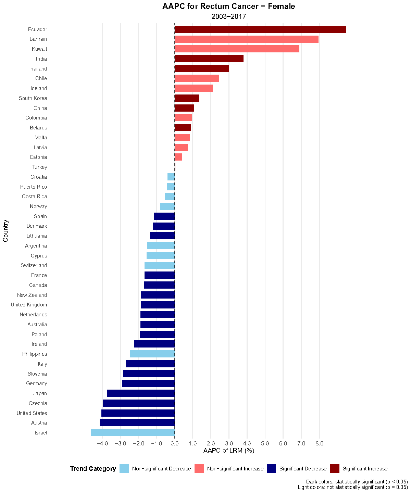

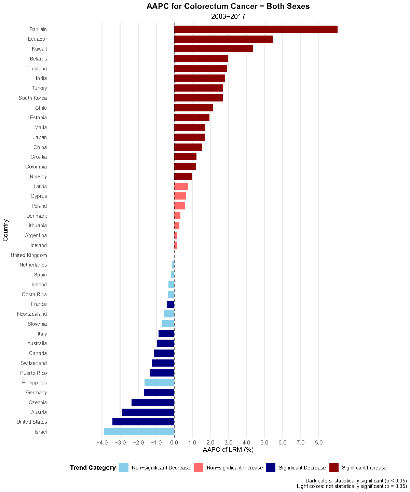

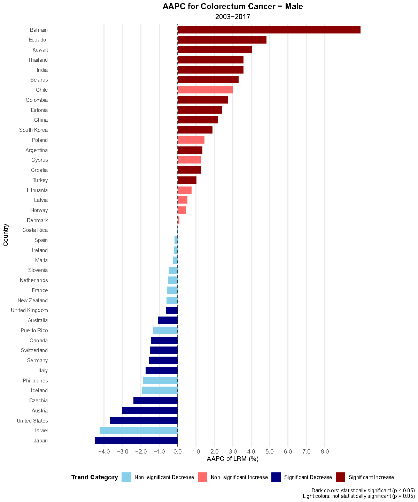

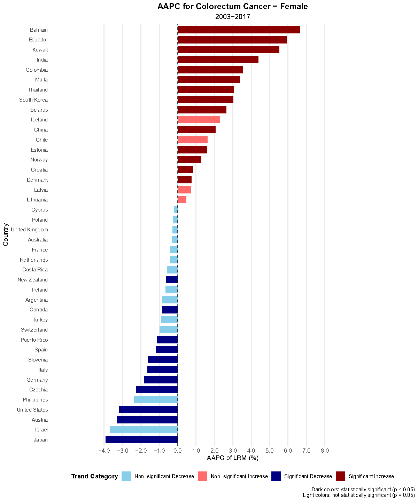
